# Cross-sectional study assessing the feasibility of measuring residents’ Quality of Life in English care homes and assessing the construct validity and internal consistency of measures completed by staff-proxy

**DOI:** 10.1101/2024.05.20.24307612

**Authors:** Ann-Marie Towers, Stacey Rand, Stephen Allan, Lucy Webster, S Palmer, Rachael E Carroll, Adam L Gordon, Gizdem Akdur, Nick Smith, Jenni Burton, Anne Killett, Barbara Hanratty, J Meyer, Karen Spilsbury, Claire Goodman

## Abstract

**Objectives:** To assess the feasibility of capturing older care home residents’ quality of life (QoL) in digital social care records (DSCRs) and the construct validity (hypothesis testing) and internal consistency (Cronbach’s Alpha) of four QoL measures.

**Design:** Cross-sectional data collected in wave one of the DACHA (**D**eveloping resources **A**nd minimum dataset for **C**are **H**omes’ **A**doption) Study, a mixed-methods pilot of a prototype minimum dataset (MDS) [1]. **Setting:** Care homes (with or without nursing) registered to provide care for older adults (>65 years) and/or those living with dementia. All homes used a DSCR system from one of two suppliers..

**Participants:** Data were extracted for 748 residents. All permanent residents, aged 65 years or older, were eligible to participate, including those lacking capacity to consent. Temporary residents and residents in their last weeks of life were excluded.

**Outcome measures and analysis:** The English language versions of: ASCOT-Proxy-Resident, ICECAP-O, EQ-5D-5L proxy and the QUALIDEM were added to the DSCRs. As there have not been any previous studies of the structural validity of the English language version of the QUALIDEM, ordinal Exploratory Factor Analysis (EFA) was applied for this measure only. Feasibility (% missing by software provider and measure), % floor/ceiling effects (>15% at lower/upper end of the scales), convergent or divergent construct validity (criterion of >75% of hypotheses accepted) and internal consistency (Cronbach’s Alpha ≥.7) were assessed for all four measures.

**Results:** The ordinal EFA of QUALIDEM did not replicate the findings of previous research. A six factors (36 items) solution was proposed and used in all subsequent analyses. There were low rates of missing data (<5%) for all items, except ASCOT-Proxy-Resident Control (5.1%) and Dignity (6.2%) and QUALIDEM item 35 (5.1%). Ceiling effects were observed for the ASCOT-Proxy-Resident and two of the QUALIDEM subscales. None of the scales had floor effects. Cronbach’s alpha indicated adequate internal consistency (α≥.70) for the ASCOT-Proxy-Resident, ICECAP-O and EQ-5D-5L proxy. There were issues with two QUALIDEM subscales. Construct validity for all measures was adequate.

**Conclusions:** The findings support the use of EQ-5D-5L, ASCOT-Proxy-Resident and the ICECAP-O in care homes for older people. Choice of measure will depend on the construct(s) of interest. More research is needed to establish the psychometric properties of the QUALIDEM in an English care home setting.

**Strengths and limitations:**

- This is the first time that quality of life measures have been piloted in routine data collection from care home digital social care records (DSCRs) in England.
- Findings support previous research that resident self-report leads to high levels of missing data. We present new evidence that collecting data through staff-proxy instead is feasible.
- Psychometric evidence supported the construct validity and internal consistency of the ASCOT-Proxy-Resident, ICECAP-O and EQ-5D-5L-Proxy.
- Missing demographic data held about residents in DSCRs meant that we were unable to describe, or assess the representativeness, of residents in the sample.
- We did not ask staff to record whether they completed the measures alone or asked the opinions of residents, family members or colleagues before making their ratings.

## INTRODUCTION

Quality of life (QoL) is an important, person-centred indicator of the quality and effectiveness of long-term social care services and support, including for older people living in care homes [2–6]. In England, there are 314,577 people living in care homes for older adults and/or dementia care [7]. Despite substantial amounts of data being held about care home residents’ health and care needs, and their use of different parts of the health and social care system, these data are not yet available in an accessible, aggregated form to inform policy, service delivery or user choice [8,9]. However, the context is changing rapidly in England, with the implementation of a data strategy for health and social care [10], aiming to drive digitalisation [11] and standardise data collected by registered social care providers, with a view to improving interoperability and to facilitate quality care delivery [12].

The DACHA Study (**D**eveloping resources **A**nd minimum dataset for **C**are **H**omes’ **A**doption) [13,14] aimed to develop and test a minimum dataset (MDS) for care homes in England. In this context, an MDS is defined as a standardised account of the demographic, social, and health characteristics and needs of older people living in long-term care (care home) settings [13]. Other countries (e.g. United States, Canada, New Zealand, and regions of the Netherlands and Belgium) have introduced or mandated MDSs for care homes. Equivalent systems have not yet been successfully adapted for the UK context [8]. Established international instruments, such as the interRAI (formerly known as the Resident Assessment Instrument) [15] were developed as a crucial tool for assessing and planning care for residents in long-term care facilities, ensuring quality care and compliance with reimbursement requirements. They historically focused on health outcomes. However, there is growing recognition of the importance of routinely capturing residents’ experiences and wellbeing [13,14,16,17] and care homes can now purchase interRAI Quality of Life (QoL) Surveys for self-report and family proxies [18].

In the UK, most care homes do not yet capture and summarise residents’ experiences and QoL in a systematic or standardised way [14]. There is also a lack of consensus around which constructs of QoL are most relevant for this population [19]. A multitude of potential instruments measure different QoL constructs, e.g. dementia, health-related and social care-related QoL [3,19]. However, relatively few QoL instruments have been developed and evaluated with the specific needs and characteristics of care home residents in mind [4]. A recent systematic review of QoL instruments used with older adults in care homes found that, of 29 instruments identified, only 14 had been psychometrically evaluated with a care home population [3]. Of these, only two, the Adult Social Care Outcomes Toolkit (ASCOT) [20] and EuroQoL-5 Dimensions EQ-5D [21], had evidence relating to their ability to detect clinically important interventional changes (responsiveness) [3]. The review also stated that no single instrument stood out as best suited to care homes for older people [3].

Based on this review and scoping of measures available, we identified ASCOT and EQ-5D and three other instruments as potentially appropriate for inclusion in the DACHA MDS: the ICEPop Capability measure for older people (ICECAP-O) [22], designed for use in economic evaluations; the DEMQOL [23], which is a measure of health-related QoL for people living with dementia; and the QUALIDEM [24], developed specifically for people with mild to severe dementia and designed for staff completion based on ratings of observable behaviour [25]. Although ICECAP-O has not yet been psychometrically assessed specifically with care home residents, it has been used with older people and another systematic review of studies reporting its psychometric properties concluded that it has good construct validity and responsiveness [26]. DEMQOL-Proxy has been widely used to measure the QoL of people living with dementia, but its psychometric properties are not as well supported as the original self-report measure [23,27,28] and concerns have been raised about the interpretation of staff proxy responses without an interviewer present [28]. A new DEMQOL-CH (care home) measure has been developed but requires further development and testing [28]. Reviews of the evidence around QUALIDEM indicate evidence of validity and reliability of the tool, but not in England [3,4].

The mode of completion (self-complete/proxy) is critical: response rates to QoL measures amongst care home residents are very low, with relatively few residents able to self-report [4,29–31] highlighting a significant methodological challenge regarding the routine measurement of QoL in care/nursing homes for older adults. Staff acting as proxies for residents can be controversial when measuring QoL, mostly due to concerns of bias [32,33], despite staff frequently collecting data about residents’ physical, psychological and social status to inform assessments and care planning [34]. Previous research has explored the level of agreement between resident and proxy-ratings using different QoL scales and, in general, the consensus is that agreement is at best ‘fair’ [35,36]. Indeed, staff sometimes worry about judging residents’ ‘subjective state’, both for psychological outcomes, such as QoL [33,37,38], and for physical outcomes, such as pain [30]. Nonetheless, use of proxy perspectives from care professionals to inform the administration of pain medication, while not considered the ‘gold standard’, is generally accepted [30].

Ideally, multi-method approaches drawing on observations and adapted qualitative interviews with residents could inform proxy-ratings and provide some information about residents’ feelings and experiences [29,30,35,39]. These methods require time and training to ensure ratings are reliable, with a degree of standardisation between individuals and services [30]. Previous research has identified a range of barriers to implementation of tools, such as lack of time and resources, and staff turnover [40]. Consequently, proxy-reporting by staff was chosen for the DACHA study to reflect a need to adopt a feasible and acceptable data collection method that would enable good coverage of data, for all residents (especially in terms of data collection burden for staff and equity for residents without close family or friends). Proxy-perspectives are not the same as self-report, they do however offer important insights into QoL of people who would otherwise be excluded [38] They have the potential to be low-burden (staff time) and easy to integrate into routine data collection using digital social care records (DSCRs), both of which are key to successful implementation of a care home MDS [40,41].

A detailed description of how the instruments were selected for inclusion in the DACHA MDS is reported elsewhere [42]. Consultations were conducted with stakeholders, including people working in and with care homes and those with lived experience (e.g. family members of residents). Four multi-item measures were chosen to represent the different QoL constructs that stakeholders told us were important to them: health-related (EQ-5D-5L-Proxy); social care-related (ASCOT-Proxy); older people’s capability wellbeing (ICECAP-O); and dementia-specific (QUALIDEM) [42]. Selection was informed by evidence of psychometric properties [3], suitability for proxy completion by staff and consideration of administrative burden (time to complete) [3]. Consultations were particularly important in selecting one dementia-specific QoL measure, with stakeholders choosing QUALIDEM over DEMQOL [42]. In response to feedback that it was important to give residents’ an opportunity to rate their own QoL, a single item QoL measure was also included: staff were asked to support residents to complete this themselves, where possible.

### AIM

To assess the feasibility of capturing residents’ QoL in DSCRs and assess the construct validity and internal consistency of the four QoL measures, collected by staff-proxy.

## METHODS AND ANALYSIS

### Study design

This analysis draws on cross-sectional data from residents’ DSCRs collected in wave one of the DACHA Study, which was a mixed-methods pilot of a prototype MDS (see study protocol for full details [1]). The study was granted ethical approval from the London Queen’s Square Research Ethics Committee (22/LO/0250).

### Public involvement in the DACHA study

Public involvement (PI) informed the design, conduct and dissemination of the DACHA study. For this study important public perspectives were taken to be those of people living in care homes, family members of people living in care homes, care workers and care home managers. A family member was a part of the team of people who developed the study and was a co-applicant for the research funding. Additionally the Public Involvement in Research Group (PiRG) at University of Hertfordshire commented on early versions of the study plan.

Throughout the study a PI team focussed on supporting public involvement and coordinating involvement with the stages of the project. The PI team was made up of the family member co-applicant, 2 academic researchers, and a Director of a care provider advocate organisation.

Involvement of care home residents was facilitated by activity providers based in care homes who met researchers online to co-produce involvement activities that would allow residents to give their opinions and perspectives to key points of the study, including data sharing, priority of different types of data for an MDS and meaning of quality of life [43]. Involvement of family members, care staff and care home managers was facilitated through an online panel, which met quarterly throughout the project. The team consulted the panel on key issues for the project in a timely way to allow the perspectives of the panel members to influence the iterative work of the project. Key issues included: priorities for an MDS; the current data environment in care homes; interpretation of findings of reviews; trusted sources of data; QoL measures; methods for recruitment and support of care homes and their residents; determining audiences for findings from the study and accessible means to communicate key messages.

### Participants

Data were extracted from the DSCRs of 748 older care home residents. Care homes (with or without nursing) were registered to provide care for older adults (>65 years) and/or those living with dementia and were located in one of three participating integrated care systems (ICSs), representing a range of geographic, socio-economic and organisational contexts [1]. ICSs are regional partnerships between NHS organisations, local government and others including third sector and social enterprises, which are responsible for co-ordinating and paying for care in England. As described in the protocol [1], all homes were using one of two DSCR systems (referred to as Provider 1 and Provider 2 hereafter). Both systems were on NHS Digital’s (now, NHS England) ‘assured solutions list’ for DSCR systems at the time of writing [44].

Within participating homes, all permanent residents aged 65 years or older were eligible to take part, including those lacking capacity to consent. Where residents were not able to consent for themselves, a nominated or personal consultee was consulted to represent their views and offer advice about participation on their behalf, as recommended by the Mental Capacity Act [45]. Residents in their last weeks of life (judged by staff) were excluded.

### Measures

Four QoL measures and a single item QoL rating scale were incorporated into the software of two DSCR providers who had agreed to participate in the study (see study protocol for full details [1]). All measures were in English.

#### Quality of life

##### Single item QoL rating scale

A single item QoL rating scale, taken from the Adult Social Care Survey in England [46] was added to software. The question asks respondents to rate their overall QoL, with responses ranging from very good (1) to very bad (7). Where possible, we asked residents to report their own QoL using this item but, where that was not possible, they could receive help or staff could answer on their behalf. To help us interpret responses, we also asked staff to tick a box indicating the type of help the resident had: no help; someone read the question to them; someone translated the question for them; someone talked through the question with them; or someone answered on their behalf (proxy).

##### ASCOT-Proxy

This is an instrument designed to measure social care-related quality of life (SCRQoL), which forms part of the ASCOT suite of measures [20,38]. It was developed for proxy completion by unpaid carers or care staff on behalf of adults using social care services, who are unable to self-report [38]. Proxy respondents are asked to rate eight questions (items) that correspond to the ASCOT SCRQoL attributes: Control over daily life, Social participation, Occupation (doing things I value and enjoy), Personal safety, Accommodation comfort and cleanliness, Personal comfort and cleanliness, Food and drink and Dignity.

Each attribute was rated according to the proxy’s own opinion (ASCOT-Proxy-Proxy) and the proxy’s view of what they think the person would say (ASCOT-Proxy-Person) against four response statements, which correspond to the ideal state, no needs, some needs and high-level needs. The dual proxy perspectives were designed to reduce any bias associated with the proxy perspective gap, i.e. differences in ratings due to proxies spontaneously adopting different approaches to proxy response [47,48], as well as, specifically for the ASCOT-Proxy, to improve acceptability of the questions to proxy respondents [38]. Based on these two proxy perspectives, the ASCOT-Proxy provides two measures of SCRQoL, the ASCOT-Proxy-Proxy and the ASCOT-Proxy-Person (here, called ASCOT-Proxy-Resident). As Preference weights for the ASCOT-Proxy instrument are not yet available we applied the weights developed for ASCOT-SCT4, which range from -.17 (worst possible) to 1 (best possible) [20].

The ASCOT-Proxy has not previously been used with care home residents as a standalone instrument. However, an adapted version for proxy-report by staff (without the use of dual proxy perspectives) is included in the care homes version (CH4) of ASCOT. This is a feasible, valid and reliable measure, with a higher % completion and better coverage than family carer proxy report [29,30]. Two recent studies of the ASCOT-Proxy completed by family carers of people with dementia, living at home, and also with care home residents using the DACHA study sample, have found that of the two measures, only the ASCOT-Proxy-Person/Resident has the same structural characteristics as the original ASCOT self-completion version (SCT4) from which the ASCOT-Proxy was adapted [49,50]. From these studies, it was concluded that the ASCOT-Proxy-Proxy perspective is still useful to include, as it may improve the instrument’s acceptability and face validity to proxy respondents, giving them an opportunity to express their own views as well as what they think the person feels [38]. However, the findings indicate that the ASCOT-Proxy-Resident should be the focus of future analyses.

##### ICECAP-O

The ICECAP-O is a measure of capability wellbeing of older adults developed for use in the economic evaluation of health and social care interventions [22]. The measure comprises five items that correspond to the following attributes: attachment, security, role, enjoyment and control [51]. UK preference weights were applied to derive a score from zero (no capability) to one (full capability) [52]. The measure has not been specifically designed or adapted for proxy report. However, it has been applied in the context of older adult care homes as a proxy-report instrument with the recommendation (pending further evidence) that it ought to be completed by professional staff, rather than family members [53].

##### EQ-5D-5L Proxy Version 2

The EQ-5D-5L is a five-level version of the EQ-5D, a measure of health-related QoL. The 5L version was developed from the original 3-level (3L) version to increase reliability and sensitivity, as well as reduce ceiling effects [21]. It includes the same five dimensions as the EQ-5D-3L, i.e., mobility, self-care, usual activities, pain/discomfort and anxiety/depression. The Proxy Version 2 of the EQ-5D-5L was designed for adults who are not able to self-report due to i.e. cognitive impairment. It asks the (proxy) respondent to rate what they think the person would say (i.e. the proxy-person perspective). In this study, the instrument was rated by care staff. Due to concerns raised about the original UK value set for the EQ-5D-5L [54], there is an ongoing UK valuation study [55]. Given this, the recommended mapping function to convert to EQ-5D-3L scores was applied, with UK values applied to generate the index score [56].

*QUALIDEM* is a measure (developed in the Netherlands) of dementia-specific QoL, based on the concept of adaptation to the perceived consequences of dementia: the original Dutch version has been validated and reported in the literature [24,25]. The questionnaire was translated into English and is available for use [25], but psychometric studies have focused on the original Dutch version or translations into German [57] or Danish [58]. The instrument comprises 40 items, which are proxy-reported by care staff on behalf of older adults with mild to severe dementia living in care homes. Of the full list of items, 37 items have previously been found to be scalable onto nine (eight strong and one weak) unidimensional subscales for people with mild to severe dementia [24,25]. Of these 37 items, 21 are suitable for people with very severe dementia that relate to six of the nine subscales [24,25]. In this study, all 40 items were included in the care home software system for completion by care staff. Each item is rated on a four-point Likert scale (never to frequently), with indicative items scoring zero for ‘never’ and contra-indicative items scoring three for ‘never’, such that higher scores always indicate better QoL in each subscales. The developers advise against calculating overall scores because subscales differ in content (between 2 and 7 items) [24].

#### Cognitive performance

Residents’ cognitive performance was one of the DACHA MDS variables identified as being important but missing from routine data collection in DSCRs [1]. Cognitive performance was measured using the Minimum Data Set Cognitive Performance Scale (MDSCPS) [59]. The scale consists of five items: dementia diagnosis, short-term memory problems, cognitive skills, ability to communicate and whether or not the person can eat and drink independently. Scores range from 0 (severe impairment)to 6 (intact cognition).

#### Functional ability

Although care notes within the DSCRs capture residents’ ability to carry out activities of daily living, they were not routinely captured in a standardised and consistent format suitable for quantitative analysis. We therefore added the Barthel Index [60] to the software, which measures the degree of assistance required with ten everyday tasks, including: feeding, bathing, grooming, dressing, continence of bowel, continence of bladder, toilet use, transfers (bed to chair and back), mobility of level surfaces and stair negotiation. Items are scored individually (0=unable to do independently; 1=needs assistance; 2= independent) and then summed and multiplied by five, to produce an overall score ranging from 0 (total dependency) to 100 (completely independent).

### Data collection

Staff completed the instruments between March and June 2023. They were instructed to complete the measures on behalf of residents, except for the single QoL item, which allowed for self-report (with or without help) or proxy report, depending on residents’ ability. Data were extracted by the software providers, in one batch (Provider 1) and four batches (Provider 2) between June and October 2023. Other health and care data pertaining to variables in the DACHA MDS (e.g. demographics, delirium, length of stay,) were also extracted (see, [1] for full description). Coded data on residents’ demographics were largely missing from DSCRs in a format suitable for quantitative analysis (despite systems being able to record this) and are therefore not reported here. Completeness of the DSCR data and the feasibility of linking it to other sources of administrative, health and care data for the purposes of populating a care home MDS, is described in full elsewhere [61].

### Statistical analysis

Complete case analyses were conducted to assess measurement properties, with the sample size for each analysis reported. First, we considered the structural validity [24] of the forty QUALIDEM items using ordinal exploratory factor analysis (EFA) on polychoric correlation matrices _[62,63]_. Ordinal EFA was applied because there have not been any previous studies of the structural validity of the English language version, against the original Dutch measure (37 items, 9 subscales) [24]. We did not conduct or report EFA for EQ-5D-5L and ICECAP-O, since they are formative measures and EFA/CFA is not appropriate [64], nor ASCOT-Proxy-Resident, since EFA and Rasch analysis has already been conducted and reported elsewhere [30]. For the ordinal EFA with QUALIDEM items, we applied Horn’s parallel analysis, using principal component analysis, without rotation, to estimate randomly generated eigenvalues in 5,000 random correlation matrixes, using the 95th percentile [65–68]. Factors were retained when the observed exceeded the random principal component eigenvalues [65,69]. When two or more factors were retained, promax rotation was applied. Items were taken to load onto a factor if the factor loading (rotated for ≥2 factors) was ≥.40 [70].

Descriptive statistics were reported for all measures (informed by the EFA for the QUALIDEM), alongside indicators of data completeness (% missing). Overall, missing data (including non-completion of whole tools) were reported to indicate the feasibility of the methods used in this study. These are reported separately by software provider because the two systems handled missing data in different ways (Provider 1 forced completion and Provider 2 did not). In both cases, there are issues with using non-completion as an indicator of feasibility, which relate to each system’s functionality^1^, which need to be considered in data interpretation. As such, feasibility of care staff completing the QoL instruments on behalf of residents was assessed by examining % missing data when at least one item in a scale had been completed.

Percentage floor (lowest score) and ceiling (highest score) was also considered for each measure, with a floor or ceiling effect indicated if reported by ≥15% of respondents [71]. For QUALIDEM, we report descriptives, completeness, floor and ceiling only for those residents rated as having ‘borderline’ to ‘severe’ cognitive impairment on the MDS CPS [59] because only six of the nine original QUALIDEM subscales are recommended for people with ‘very severe impairment’ [24]. Since there were only n=79 residents rated ‘very severe’ on the MDS CPS, we were unable to run the analysis for these cases separately.

Construct (convergent or divergent) validity of the QoL measures was assessed by hypothesis testing about expected relationships with other outcomes measures, using Spearman rank correlation (p-value less than 0.01). Correlation coefficients were interpreted as weak (<.3), moderate (.3 to .5) or strong (>.5) [72]. These hypotheses were based on previous studies using the ASCOT-Proxy or other ASCOT measures (SCT4, CH4), or developed *a priori* based on the measurement constructs (see **Table 5**). A criterion of >75% of hypotheses accepted was considered as sufficient evidence of construct validity [73].

Internal consistency was considered using Cronbach’s alpha, with a value of ≥.7 taken to be acceptable [74]. COSMIN reporting guidance advises that an assessment of internal consistency is not required for formative measures [64]. Preference-based measures (EQ-5D-5L, ASCOT and ICECAP) are generally accepted to be formative [75], however for comparability with previous research [26,29,35], we have assessed internal consistency in this study.

We used the COSMIN Study Design Checklist rule of thumb for adequacy of sample size for EFA, internal consistency and construct validity by hypothesis testing. In all cases, >100 participants is ‘very good’ [64].

All analyses were conducted in STATA 16 [76].

## RESULTS

### Structural validity of QUALIDEM

This is reported first because the findings inform other analyses and reporting. The ordinal EFA of QUALIDEM did not replicate the nine-factor structure proposed by the original developers, i.e. 37 items relating to nine subscales of dementia-related QoL [24,25]. First, we had to omit two items (33-Criticizes the daily routine and 37 - Indicates feeling worthless) due to linear dependencies that led to indefinite matrices when conducting ordinal EFA. With the remaining 38 items, Horn’s parallel analysis indicated a six-factor solution, for which 36 items loaded onto at least one of the six factors with loading ≥.40 (see **Table 1)**. These six factors (36 items) related to positive and negative affect (including mood and behaviour) (Subscale 1. 15 items), restlessness, tension and agitation (Subscale 2. 5 items), enjoyment of meals/food (Subscale 3. 2 items), boredom and disengagement (Subscale 4. 6 items), social engagement (Subscale 5. 5 items) and anxiety or low mood (Subscale 6. 3 items). Items 17 and 26 did not load on to any of these six factors. This six-subscale (36 item) solution is used in all subsequent analyses.

**Table 1.** Exploratory Factor Analysis of QUALIDEM (n=540)

|  | Factor 1<br>loadings | Factor 2<br>loadings | Factor 3<br>loadings | Factor 4<br>loadings | Factor 5<br>loadings | Factor 6<br>loadings | Uniqueness |
| --- | --- | --- | --- | --- | --- | --- | --- |
| 1. Is cheerful | .93 |  |  |  |  |  | 0.19 |
| 2. Makes restless movements |  | .93 |  |  |  |  | 0.30 |
| 3. Has contact with other residents | (.51) |  |  |  | .54 |  | 0.35 |
| 4. Rejects help from nursing assistants | .53 |  |  |  |  |  | 0.21 |
| 5. Radiates satisfaction | .77 |  |  |  |  |  | 0.44 |
| 6. Makes an anxious impression |  |  |  |  |  | .73 | 0.30 |
| 7. Is angry | .47 | (.45) |  |  |  |  | 0.25 |
| 8. Is capable of enjoying things in daily life | .65 |  |  |  |  |  | 0.27 |
| 9. Does not want to eat |  |  | .93 |  |  |  | 0.15 |
| 10. Is in a good mood | .91 |  |  |  |  |  | 0.19 |
| 11. Is sad |  |  |  |  |  | .70 | 0.27 |
| 12. Responds positively when approached | .81 |  |  |  |  |  | 0.15 |
| 13. Indicates that he or she is bored |  |  |  | .60 |  |  | 0.52 |
| 14. Has conflicts with nursing assistants | .52 |  |  |  |  |  | 0.18 |
| 15. Enjoys meals |  |  | .91 |  |  |  | 0.15 |
| 16. Is rejected by other residents |  | .67 |  |  |  |  | 0.41 |
| 17. Accuses others |  |  |  |  |  |  | 0.38 |
| 18. Takes care of other residents |  |  |  |  | .87 |  | 0.25 |
| 19. Is restless |  | .98 |  |  |  |  | 0.19 |
| 20. Openly rejects contact with others | .43 |  |  |  |  |  | 0.33 |
| 21. Has a smile around the mouth | .88 |  |  |  |  |  | 0.24 |
| 22. Has tense body language |  | .51 |  |  |  |  | 0.45 |
| 23. Cries |  |  |  |  |  | .50 | 0.48 |
| 24. Appreciates help he or she receives | .71 |  |  |  |  |  | 0.17 |
| 25. Cuts himself/herself off from environment | (.42) |  |  | .59 |  |  | 0.40 |
| 26. Finds things to do without help from others |  |  |  |  |  |  | **0.60** |
| 27. Indicates he or she would like more help |  |  |  | .83 |  |  | 0.40 |
| 28. Indicates feeling locked up |  |  |  | .41 |  |  | 0.41 |
| 29. Is on friendly terms with one or more |  |  |  |  | .61 |  | 0.24 |
| 30. Likes to lie down |  |  |  | .48 | (.42) |  | 0.59 |
| 31. Accepts help | .57 |  |  |  |  |  | 0.30 |
| 32. Calls out |  |  |  |  | .40 |  | 0.42 |
| 33. Criticizes the daily routine (omitted) |  |  |  |  |  |  | Omitted |
| 34. Feels at ease in the company of others | .67 |  |  |  |  |  | 0.41 |
| 35. Indicates not being able to do anything |  |  |  | .66 |  |  | 0.40 |
| 36. Feels at home on the ward | .74 |  |  |  |  |  | 0.46 |
| 37. Indicates feeling worthless (omitted) |  |  |  |  |  |  | Omitted |
| 38. Enjoys helping with chores on the ward |  |  |  |  | .51 |  | **0.71** |
| 39. Wants to get off the ward |  | .44 |  |  |  |  | 0.41 |
| 40. Mood can be influenced in positive sense | .74 |  |  |  |  |  | 0.41 |

Only factors loading $\geq .40$ are reported. Items with uniqueness $\geq .60$ shown in **bold**

### Feasibility

Missing data are reported in **Table 2**. Overall, % missing data were higher for Provider 2, compared to Provider 1. Differences in the two software systems for providers - specifically, Provider 1 required forced completion, but Provider 2 did not - were likely to have partly influenced variations in data completion. It may also have been affected by the longer period between consent and data completion for Provider 2 due to delays in finalising and releasing the instruments to care homes: due to participants no longer being resident (i.e., due to hospitalization or death). This is supported by the higher % of residents where no data were completed for Provider 2, compared to Provider 1.

**Table 2.** % missing data.

|  | Overall %<br>missing data<br>software provider<br>1 † | Overall %<br>missing data<br>software provider<br>2 † | Non-completion<br>of scale % of<br>sample<br>Provider 1 † | Non-completion<br>of scale % of<br>sample<br>Provider 2 † | % missing data<br><5% for all items<br>where ≥1 items<br>completed? |
| --- | --- | --- | --- | --- | --- |
| ASCOT-Proxy-Resident | 15.3% | 37.9% | 9.4% | 24.2% | **No**<sup>1</sup> |
| ICECAP-O | 8.8% | 26.0% | 8.8% | 25.4% | Yes |
| EQ-5D-5L Proxy 2 | 7.7% | 14.7% | 7.6% | 12.3% | Yes |
| QUALIDEM 1: Positive or negative affect | 91.6%* | 22.8% | 11.2% | 13.7% | **No**<sup>1</sup> |
| QUALIDEM 2: Restlessness, tension and agitation | 12.6% | 18.6% | 11.2% | 13.7% | Yes |
| QUALIDEM 3: Enjoys meals/food | 12.6% | 20.7% | 11.2% | 14.4% | Yes |
| QUALIDEM 4: Boredom and disengagement | 15.4% | 26.6% | 11.2% | 13.7% | **No**<sup>1</sup> |
| QUALIDEM 5: Social engagement | 12.6% | 18.8% | 11.2% | 13.7% | Yes |
| QUALIDEM 6: Anxiety and low mood | 12.6% | 16.9% | 11.2% | 13.7% | Yes |
| ASCS Overall QoL (single item) | 8.2% | 20.9% | n/a | n/a | n/a |

**Bold text** indicates that the criterion for psychometric evaluation was not met.
† Overall sample $n=748$ Provider 1 $n=170$ Provider 2 $n=578$
For QUALIDEM only, we excluded residents with very severe dementia on the Minimum Data Set Cognitive Performance Scale (MDS CPS) ( $n=79$ ) to leave an overall sample of $n=669$ split between provider 1 $n=143$ and provider 2 $n=526$ .
<sup>1</sup> Full details reported in Table 3.
\*Due to missing item in the software provider system.

Due to these data limitations, the feasibility of care staff completing the QoL instruments on behalf of residents was assessed by examining % missing data when at least one item in a scale had been completed (see **Tables 2** and **3**). Apart from QUALIDEM item 1, which was omitted in the first release of the software to care homes by Provider 1, none of the QoL items had % missing data ≥7%. There were low rates of missing data (<5%) for all items in the multi-item QoL instruments, except ASCOT-Proxy-Resident Control (5.1%) and Dignity (6.2%) and QUALIDEM item 35 (5.1%) Overall, this indicates that the multi-item QoL instruments were feasible for care home staff to complete.

Of the n=613 cases (81%) where the single item ASCS QoL item was completed, 14.9% (n=91) were completed by the resident without help and 27.8% (n=170) were completed by staff proxy, without any involvement of the resident. The remaining responses, except one case of missing data, (57.3%, n=351) were completed by the resident with assistance from care staff, e.g. to read, talk through and/or translate questions.

### Floor and ceiling effects

Descriptive statistics and summary of the psychometrics, including % floor/ceiling, are reported in **Table 4**. Social care-related QoL, measured by the ASCOT-Proxy-Resident, was higher than expected with a mean of 0.83 and a ceiling effect, i.e., >15% at upper end of the scale. Previous research using ASCOT in care homes has used the mixed-methods tool (ASCOT-CH4), in which trained researchers rate residents’ SCRQoL after conducting structured observations, staff interviews and speaking to residents [29,77]. In studies using this method of data collection, residents’ mean SCRQoL scores ranged from 0.74-0.77 [30].

**Table 3.** Missing data by item, where ≥1 item completed.

| **Measure** | **% Missing** |
| --- | --- |
| **ASCOT-Proxy-Resident (n=592)** |  |
| 1. Food & drink | 3.4% |
| 2. Home comfort & clean | 3.4% |
| 3. Personal comfort & clean | 2.2% |
| 4. Social participation | 3.9% |
| 5. Occupation | 4.9% |
| 6. Control over daily life | **5.1%** |
| 7. Personal safety | 3.2% |
| 8. Dignity | **6.8%** |
| **QUALIDEM 1. Positive or negative affect (n=574)** |  |
| 1. Is cheerful | **19.9%<sup>1</sup>** |
| ... All other items | <2% |
| **QUALIDEM 4. Boredom and disengagement (n=574)** |  |
| 13. Indicates he or she is bored | 3.3% |
| 25. Cuts him/herself off of environment | <2% |
| 27. Indicates he or she would like more help | 2.6% |
| 28. Indicates feeling locked up | 3.5% |
| 30. Likes to lie down | 2.1% |
| 35. Indicates not being able to do anything | **5.1%** |

<sup>1</sup> Due to omission of item in first release of the measure in software provider 1's system. If those cases affected by this error are not considered, the % missing is <2%.

**Table 4.** Descriptive Statistics and Internal Consistency.

|  | Mean, Std. Dev | Range | N †† | % floor | % ceiling | Cronbach's α (no. of items) |
| --- | --- | --- | --- | --- | --- | --- |
| ASCOT-Proxy-Resident † | .83, .19 | -.17 to 1.00 | 503 | <2% | **17.7%** | .83 (8) |
| ICECAP-O | .73, .21 | 0 to 1 | 583 | <2% | 3.4% | .81 (5) |
| EQ-5D-5L Proxy 2 | .33, .35 | -.59 to 1 | 650 | <2% | <2% | .74 (5) |
| QUALIDEM 1: Positive or negative affect | 35.52, 7.50 | 11 to 45 | 418 | 0% | 7.4% | .92 (15) |
| QUALIDEM 2: Restlessness, tension and agitation | 10.87, 3.42 | 1 to 15 | 553 | 0% | **19.7%** | .78 (5) |
| QUALIDEM 3: Enjoys meals/food | 4.65, 1.37 | 0 to 6 | 542 | <2% | **38.8%** | .72 (2) |
| QUALIDEM 4: Boredom and disengagement | 12.24, 3.58 | 1 to 18 | 507 | 0% | 6.7% | **.66 (6)** |
| QUALIDEM 5: Social engagement | 7.78, 3.44 | 0 to 15 | 552 | <2% | <2% | .72 (5) |
| QUALIDEM 6: Anxiety and low mood | 5.65, 2.12 | 0 to 9 | 562 | <2% | 9.3% | .75 (3) |
| ASCS Overall QoL item from best (1) to worst (7) | 3.16, 1.08 | 1 to 7 | 613 | n/a | n/a | n/a |
| Barthel Index from lowest (0) to highest (100) independence | 41.49, 30.19 | 0 to 100 | 630 | n/a | n/a | n/a |
| MDS CPS from very severe impairment (0) to intact (6) | 3.10, 2.01 | 0 to 6 | 582 | n/a | n/a | n/a |

**Bold text** indicates that the criterion for psychometric evaluation was not met.
† The descriptive statistics and psychometrics are only reported further for the ASCOT-Proxy-Resident, using preference weights developed for the ASCOT-SCT4 (Netten et al, 2012). For further discussion and justification of our focus on ASCOT-Proxy-Resident, not –Proxy-Proxy (see Rand et al, 2024; Silarova et al, 2023).
†† Overall sample $n=748$ Provider 1 $n=170$ Provider 2 $n=578$
For QUALIDEM only, we excluded residents with very severe dementia on the Minimum Data Set Cognitive Performance Scale (MDS CPS) ( $n=79$ ) to leave an overall sample of $n=669$ split between provider 1 $n=143$ and provider 2 $n=526$ .

There were no floor or ceiling effects for the ICECAP-O, measuring capability wellbeing. We cannot compare mean scores with previous research because ICECAP-O has not previously been used in British care homes. However, the DACHA sample had a lower proxy-reported mean QoL score compared to a community sample of older people (>65 years) in England (0.73 vs 0.81), which is consistent with differences in the functional ability of the two samples [78].

Two of the QUALIDEM subscales had ceiling effects (2, 3) with >15% at the upper end of the scale. The subscales we identified in this study differ from the subscales identified by the developers based on the original Dutch version [22], as applied also to the Danish version [55], or German translations [54]. Therefore, there is no comparable data on subscales means.

Staff used the full scale to capture residents’ health-related QoL using the EQ-5D-5L proxy with less than 2% of scores at the top and bottom of the range. There was a mean score of 0.33, which is in line with previous research [29].

Mean Barthel and cognitive performance scores were as expected for this population based on previous research, and indicate severe dependency [77,79]. Although we do not have demographic information, these are reassuring indicators of the representativeness of the sample to the care home population of each ICS [40].

### Internal consistency

Cronbach’s alpha indicated adequate internal consistency (α≥.70) for the ASCOT-Proxy-Resident, ICECAP-O, EQ-5D-5L proxy and QUALIDEM, except for QUALIDEM Subscale 4 (boredom and disengagement) based on the EFA conducted for this study (α≤.70, **Table 1**). QUALIDEM Subscale 1

(positive and negative affect) also had very high internal consistency (α≥.90), which may indicate redundancy of items.

### Construct validity

The construct validity analysis by hypothesis testing is reported in Table 5. As >75% of the proposed hypotheses were accepted for each set of hypotheses, there is evidence of adequate construct validity.

**Table 5.**
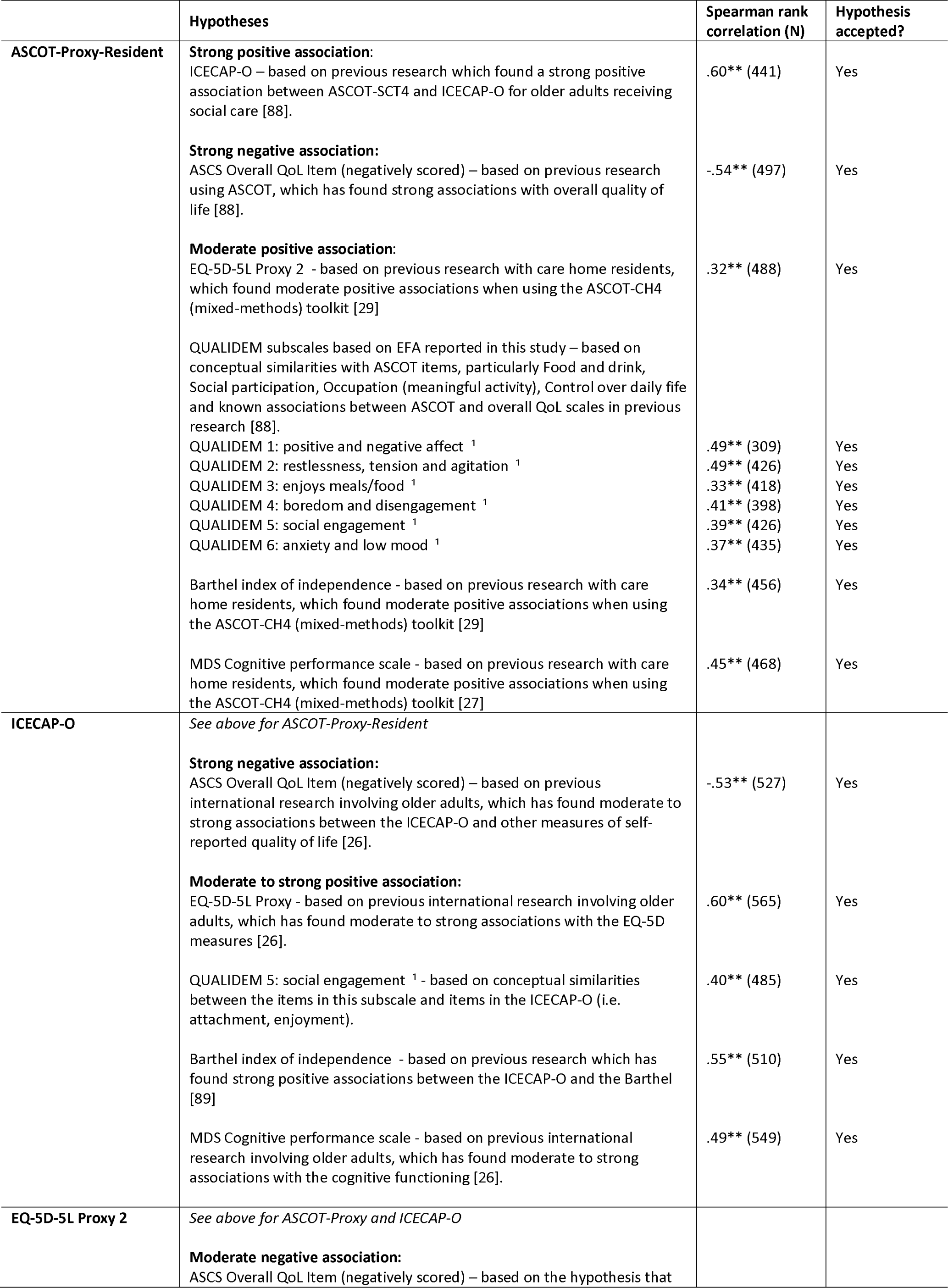
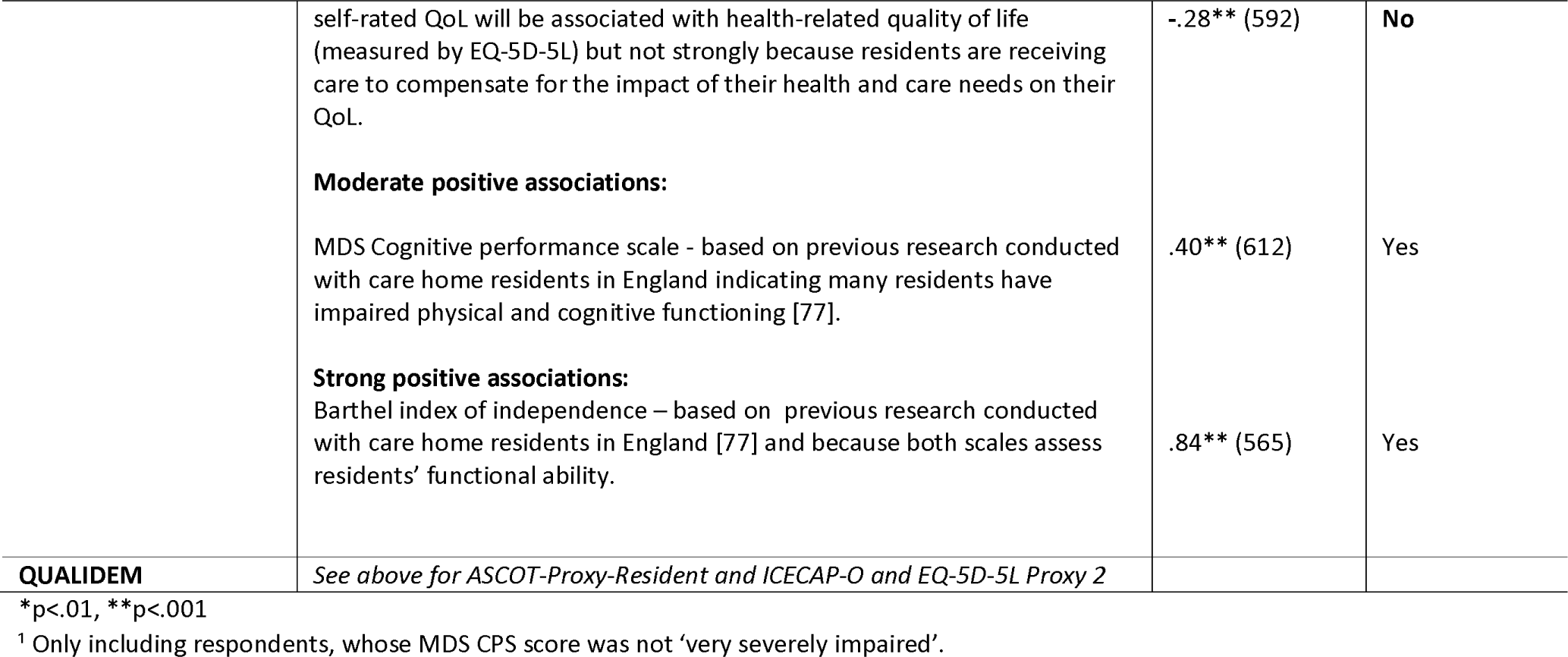
Construct validity by hypothesis testing.

## DISCUSSION

This study sought to explore the feasibility of routinely capturing QoL data about care home residents and assessed the construct validity and internal consistency of four QoL measures, completed by staff proxies. The measures were integrated into two DSCR systems, both of which were on the NHS Digital ‘assured solutions’ list, yet the two systems differed in their tolerance of missing data and how they implemented the measures in participating homes. Forced completion of the items within the measures led to less missing data overall. Delays finalising and releasing the instruments to care homes for Provider 2, led to a longer gap between resident recruitment and completion of the measures and reduced the time staff had to complete the measures before data extraction. This also contributed to higher rates of non-completion, due to participants no longer being resident in the care home (i.e., due to hospitalization or death).

Implementation issues aside, once staff began to complete the QoL measures they were likely to finish them, indicating completion by staff proxy is a feasible method of collecting QoL data for the purposes of a care home MDS. Only the ASCOT-Proxy-Resident Dignity item had more than 6% missing data. This item is important when capturing the impact of social care on people’s QoL [20] and was acceptable during the development of the ASCOT-Proxy [38], yet staff and family proxies alike appear to find this more difficult to judge than the other domains [38,50]. ASCOT-Proxy-Resident Dignity asks the proxy to rate the effect of help from paid carers on how the resident thinks and feels about themselves (from the resident’s perspective). This involves several empathetic perspective shifts, which proxies may find difficult to navigate cognitively and/or judge through their day-to-day interactions with the person. Qualitative interviews and focus groups with staff exploring their experiences of completing the measures have been undertaken and will be reported separately [1].

Previous reviews have proposed the QUALIDEM as among the best QoL measures for use in data collection in care homes for older people [4,80]. It was the dementia QoL scale that achieved the most support from stakeholders for the DACHA study, hence its inclusion in the study [42]. However, the mixed nature of prior evidence of its psychometric properties have been noted [80]. The developers of the original measure, in Dutch, indicated some issues with scalability and internal reliability, for some subscales; furthermore, assessment of the structural validity of the German translation did not support the original subscales [57,81]. Here, we present the first EFA on the English tool, which indicated a six-factor solution, using 36 of the original 40 items. These do not correspond to the original Dutch or German translation subscales [25,81]. There were also issues with the internal consistency for two of the subscales: boredom and disengagement (subscale 4), which did not meet the criteria (α≤.70, **Table 1**) and positive and negative affect (subscale 1), which had very high internal consistency (α≥.90), potentially indicating redundancy of items. Despite adequate construct validity, the mixed evidence for structural validity and internal consistency, both in this study and previous studies, means that we are not able to recommend the inclusion of QUALIDEM in a UK care home MDS at this time. Future research should explore the replicability of these findings in the English language.

Overall, the psychometric evidence (internal consistency, construct validity and also, structural validity, where appropriate) supported the use of the other three multi-item measures. These were also the measures that had the best psychometric evidence when considering the measures to include in the DACHA MDS, as well as alignment to the constructs of (social/long-term) care-related and health-related QoL that are most useful in reflecting on the quality and effectiveness of care delivered in the care home context [3]. The ASCOT-Proxy-Resident had a ceiling effect, but this is common and reflects the fact that ASCOT captures the impact of social care on QoL – if good quality care is being delivered and meeting people’s needs and preferences, they will score highly, and this is a desirable state. This is supported by the findings of previous research in care homes showing a positive association between residents’ SCRQoL and care home quality ratings [30,82,83].

Most residents required help (e.g. to read or talk through the question) to complete the single-item QoL scale, with over a quarter completed by staff-proxy with no resident input at all. Only 15% of residents in this study completed this question without any help at all. This is in line with previous research in English care homes for older adults, which found that less than 25% of residents could give their views of their own care-related QoL using a structured ASCOT questionnaire, whereas around 60% could talk about the care-related QoL if questions were asked in a flexible, qualitative interview [77]. It is likely that if residents had self-completed the longer QoL measures in DACHA, we would have had substantial missing data, affecting the ability to generate overall scores and interpret the results.

A limitation of this study is that, despite expecting staff to complete the measures by proxy, we cannot be sure of the extent to which staff discussed the questions with residents before/while completing them [84]. We only collected this information for the single item QoL question. Ideally, in future data collection, detail on exactly how proxies completed the measure (i.e. on their own, after speaking with the resident, or by asking the resident to give their own view) should be captured and considered in analyses. Another limitation of this study is that most care homes did not complete the demographic fields in the DSCRs. Consequently, information about gender, ethnicity, age and other demographic data were missing from the data extraction. However, the psychometric analysis reported in this paper did not require these data and further analysis using the data to better understand the quality of life of care home residents has used the complete DACHA MDS [61], in which demographic data has been populated through linkage with NHS data [61,85]. The linked data has been compared to the overall care home resident population in England to explore representativeness: findings indicate that the DACHA MDS sample is comparable by sex and type of care home but the very old and ‘White’ ethnic group are over-represented [86].

For DSCR data to be consistently used to populate a care home MDS, greater standardisation of the approach to missing data should be considered. Nonetheless, the evidence reported here indicates that it is feasible to routinely capture data about residents’ QoL through staff-proxies. The study has demonstrated that it is not feasible to consistently collect data from care home residents through self-report alone. Most residents will require help in the form of reading the questions, talking through the responses and marking the answers. A substantial proportion would be excluded entirely without using proxy-report. Three of the four QoL measures piloted had good psychometric properties for internal consistency and construct validity by hypothesis testing: the EQ-5D-5L (health-related QoL), the ASCOT-Proxy-Resident (social care-related QoL) and the ICECAP-O (capability wellbeing). As a key purpose of measuring resident QoL is to assess care quality and effectiveness; it is vital that the QoL measures included in the MDS are responsive to the quality, safety and effectiveness of care. This should be explored in future research.

This study is the first to pilot the inclusion of QoL measures in DSCRs in England. It was not possible to make specific recommendations about which of the three QoL measures with satisfactory performance should be prioritised for inclusion in an MDS. Each measures a different QoL construct and, as such, further work would be required with key stakeholders, if a choice was required. Multiple QoL measures add to question burden in an MDS but, uniquely, bring person-centred outcomes to otherwise largely clinical and process-oriented datasets. There may be a strong case for including more than one. Staff were not given training or detailed guidance beforehand, only the written instructions already included by the authors of the scales. Despite this, most measures were completed in full once staff made a start. The ASCOT-Proxy-Resident had slightly higher levels of missing data for some items (e.g. dignity). This may also indicate that staff would benefit from more guidance or support to interpret and complete these items as part of routine care. Ongoing work to support the use of ASCOT in care planning in care homes in Sweden [87] indicates that these issues can be addressed by training key members of staff to be QoL champions, mentoring other staff. The care planning approach, which involves conversations with residents and family members, also better integrates QoL into routine care by identifying how practice will maintain or improve QoL. This is one of the core principles of the DACHA MDS [14] and may be useful when considering the implementation of QoL measures in DSCRs in the future.

## Funding statement

This project is funded by the National Institute for Health Research (NIHR) Health Service Research and Delivery programme (HS&DR NIHR127234) and supported by the NIHR Applied Research Collaboration (ARC) East of England.

AMT, ALG, BH, KS, G_P,_ AK, and CG are supported by the NIHR Applied Research Collaborations in Kent, Surrey and Sussex; East Midlands; North East and North Cumbria; Yorkshire and Humber and East of England respectively.

CG, ALG and KS are NIHR Senior Investigators.

The views expressed are those of the authors and not necessarily those of the NIHR or the Department of Health and Social Care.

## Data Availability

Pseudonymised data will be available on request from the corresponding author, AMT, following a 24 month embargo from the date of publication.

## Acknowledgments

We would like to acknowledge and thank the Public Involvement and Engagement Panel members, software providers, care homes, care staff and residents for participating in the study, as well as the wider DACHA study team whose work informed the development of the DACHA (**D**eveloping resources **A**nd minimum dataset for **C**are **H**omes’ **A**doption) Minimum Data Set and contributed to the selection of the quality of life measures included in the pilot.

## Footnotes

1 Specifically: **Provider 1** ’s system required forced completion for items in ICECAP-O, QUALIDEM, EQ-5D-5L; for ASCOT-Proxy only, it was possible to select ‘don’t know’, coded as missing data. For **Provider 2**, the system did not require completion of all items. There was no user prompt if items were not completed. Therefore, it was possible to only partially complete each measure, due to either deliberate non-completion (i.e. due to item acceptability or feasibility) or user error of omission.

